# SARS-CoV-2 (COVID-19) infection in pregnant women: characterization of symptoms and syndromes predictive of disease and severity through real-time, remote participatory epidemiology

**DOI:** 10.1101/2020.08.17.20161760

**Authors:** Erika Molteni, Christina M. Astley, Wenjie Ma, Carole H Sudre, Laura A. Magee, Benjamin Murray, Tove Fall, Maria F. Gomez, Neli Tsereteli, Paul W. Franks, John S. Brownstein, Richard Davies, Jonathan Wolf, Tim D Spector, Sebastien Ourselin, Claire J Steves, Andrew T Chan, Marc Modat

## Abstract

**Objective:** To test whether pregnant and non-pregnant women differ in COVID-19 symptom profile and severity. To extend previous investigations on hospitalized pregnant women to those who did not require hospitalization.

**Design:** Observational study prospectively collecting longitudinal (smartphone application interface) and cross-sectional (web-based survey) data.

**Setting:** Community-based self-participatory citizen surveillance in the United Kingdom, Sweden and the United States of America.

**Population:** Two female community-based cohorts aged 18-44 years. The discovery cohort was drawn from 1,170,315 UK, Sweden and USA women (79 pregnant tested positive) who self-reported status and symptoms longitudinally via smartphone. The replication cohort included 1,344,966 USA women (134 pregnant tested positive) who provided cross-sectional self-reports.

**Methods:** Pregnant and non-pregnant were compared for frequencies of symptoms and events, including SARS-CoV-2 testing and hospitalization rates. Multivariable regression was used to investigate symptoms severity and comorbidity effects.

**Results:** Pregnant and non-pregnant women positive for SARS-CoV-2 infection were not different in syndromic severity. Pregnant were more likely to have received testing than non-pregnant, despite reporting fewer symptoms. Pre-existing lung disease was most closely associated with the syndromic severity in pregnant hospitalized women. Heart and kidney diseases and diabetes increased risk. The most frequent symptoms among all non-hospitalized women were anosmia [63% pregnant, 92% non-pregnant] and headache [72%, 62%]. Cardiopulmonary symptoms, including persistent cough [80%] and chest pain [73%], were more frequent among pregnant women who were hospitalized.

**Conclusions:** Symptom characteristics and severity were comparable among pregnant and non-pregnant women, except for gastrointestinal symptoms. Consistent with observations in non-pregnant populations, lung disease and diabetes were associated with increased risk of more severe SARS-CoV-2 infection during pregnancy.

**Tweetable abstract:** Pregnancy with SARS-CoV-2 has no higher risk of severe symptoms. Underlying lung disease or cardiac condition can increase risk.

## 1. Introduction

The COVID-19 pandemic is caused by the SARS-CoV-2, a newly identified enveloped RNA-β-coronavirus ^1,2^. Early on, pregnant women were regarded as vulnerable group, at greater risk of severe morbidity and mortality, based on previous studies of smaller coronavirus outbreaks, and the theoretical risks associated with immunosuppression of pregnancy ^3–5^. However, substantial literature has now documented that, among hospitalized pregnant women, antecedent symptoms and risk factors for severe disease are similar to those outside pregnancy ^6^, and few hospitalized pregnant women require admission to intensive care or intubation, although preterm birth, Caesarean delivery, and stillbirth may be increased compared with women without COVID-19, and vertical transmission possible (86 studies to 8 Jun 2020) ^7–10^. SARS-CoV-2 positive patients develop dry cough, fever, dyspnea, fatigue and bilateral lung infiltrates on imaging in the severe cases ^11^. Hospitalized pregnant women positive for SARS-CoV-2 manifest similar symptoms ^7,12,13^. However, little is known about pregnant women affected by SARS-CoV-2 infection in the community, many of whom recover without hospitalization ^14^.

Smartphone and web-based applications for population-based syndromic surveillance are citizen science tools that can facilitate rapid acquisition of extensive epidemiological data as a pandemic evolves ^15^. These data can inform public-health policies, enhance the speed of the healthcare response, shape the community services, and alert the general population to urgent health threats ^16^. Smartphone applications (apps) were used prior to the COVID-19 pandemic to remotely advise on prenatal health, and maternal health behaviours, including gestational weight gain and smoking cessation ^17^. Many eHealth initiatives were launched at the onset of the pandemic, with most using single, cross-sectional reporting methods to inform SARS-CoV-2 epidemiology ^18^. We present findings from a unique, longitudinal community-based symptom-tracking system that identified both test positive and suspected (but untested) SARS-CoV-2 infected pregnant women, who were followed prospectively to assess the need for hospitalization. Furthermore, we replicated key findings, using an independent, cross-sectional symptom survey.

We present data from a cohort of women of childbearing age, including pregnant women who report test-positive SARS-CoV-2. Despite presenting a wide spectrum of disease manifestations, these pregnant women rarely required hospitalization. In order to include non-tested subjects who developed symptoms during the onset of the pandemic, when testing resources were still limited, we developed a model to predict positivity to SARS-CoV-2 based on symptoms, specific to female population in childbearing age. We sought to characterize the differences in the SARS-CoV-2 symptom profiles and severity between pregnant and non-pregnant women who did and did not receive hospitalization. We identified demographic characteristics and comorbidities that modified symptom severity of SARS-CoV-2 in pregnancy.

## 2. Materials and methods

### 2.1 Study Populations

We developed a symptom-based prediction method to identify suspected COVID-19 cases among women 18-44 years of age from a discovery cohort. Results were replicated in an independent, cross-sectional cohort with different survey methodology.

#### Discovery Cohort

The COVID Symptom Study smartphone-based application (app), developed by Zoe Global Limited, and having almost four million users from the general population in UK, 280,000 from USA and around 175,000 from Sweden. Users self-report daily information about their overall health status, as well as their symptoms (from a pre-defined list, to standardise input) ^19, 20^. We included all pre-menopausal (if menopausal status was reported) women aged 18 to 44 years, who used the app between 24 March and 7 June 2020, and specified their pregnancy status at baseline (pregnant or not pregnant) included symptom profiles, outcomes on testing positive for SARS-CoV-2, and hospitalization (Supplementary Material 1).

#### Replication Cohort

The Facebook COVID-19 Symptom survey, launched in the USA and hosted by the Carnegie Mellon Delphi Research Center. Surveys were conducted using sampling strategies and survey weights to ensure respondents were representative of the USA source population ^21^ (Supplementary Material 1). Using data from launch (6 April 2020) through 7 June 2020, we identified surveys from 1,344,966 female respondents who indicated their pregnancy status and age 18-44 years ^22^. Users specified if they had experienced specific symptoms over the last 24 hours, in addition to answering demographic and infection-related questions.

### 2.2 Pregnancy groups, symptoms, syndromes and outcomes

#### Pregnancy status

Women were divided into pregnant and non-pregnant subgroups, based on self-reported pregnancy status, ascertained once near the start of follow-up in the discovery cohort, and at each survey for the replication cohort. Gestational age, at the time pregnancy was ascertained, was available only for the discovery cohort.

#### COVID-19 Test and Suspected Positive

Self-reported COVID-19 testing was used to identify women with SARS-CoV-2 infection (termed *test positive*). Test positives were considered *symptomatic positive* if they reported at least one of the tracked symptoms. The type of test (e.g. PCR, serology) was not ascertained, and those reporting a pending test were excluded.

Suspected positive cases were imputed, based on a previously published method for the computation of a test-positive prediction score ^20^. The model was retrained for pregnancy age distribution, based on a bootstrapped train-test scheme in the discovery cohort, and using a strict mapping to equate symptoms ascertained in both the discovery and replication cohorts. We defined the outcome of suspected COVID-19 (termed *suspected positive*) for anyone with a score-based imputation probability above a computed threshold (Supplementary Material 2).

#### Hospitalization and Syndrome Severity

Individuals were considered to have been hospitalized when they indicated being either admitted to or discharged from hospital in their daily reporting, within one week before/after reporting at least one of the tracked symptoms. Symptoms, test results and hospitalization can be reported anytime and with no interdependencies in the app, and symptom reporting is not censored after input of test results. Symptom severity was thus defined as the weighted sum of symptoms based on peak presentation when comparing individuals reporting hospital visit with individuals who did not, in the training set of the discovery cohort (Supplementary Material 3). Symptoms were equated in the two cohorts. The weighting was then normalized so that the severity index ranges from 0 (no symptom) to 1 (all symptoms).

### 2.4 Statistical analysis

A power analysis was conducted to assess the suitability of the samples size. To account for the difference in age distributions between pregnant and non-pregnant groups, age-standardization was performed, by calculating weights for the non-pregnant women, to standardize to the age-distribution of the pregnant population (Supplementary materials 4 and 5).

#### Symptoms

To explore differences in the symptom profile between pregnant and non-pregnant women who tested or were suspected positive for SARS-CoV-2 and who also required hospitalization or sought care, we applied univariate unconditional age-weighted logistic regression for each of 18 symptoms ascertained in either the discovery cohort, the replication or in both. We then conducted multivariate analysis on symptoms grouped into clusters by body system, as shown in Table 2, and normalized to range from 0 to 1.

#### Severity of syndrome

To assess symptom severity differences between pregnant and non-pregnant women who tested or were suspected positive for SARS-CoV-2 infection and were hospitalized, univariate unconditional age-weighted regression was applied to the pregnant and non-pregnant groups of the discovery cohort, with the severity index as a response variable. The analysis was repeated for this cohort among those who reported to have been ‘seen at a hospital for their symptoms’, conditional on those who predicted or tested positive for SARS-CoV-2.

#### Hospitalization

To explore differences in the symptom profiles between hospitalized and non-hospitalized pregnant women positive for SARS-CoV-2, the frequency and percentage of women reporting each symptom were calculated for the discovery cohort. Symptoms were ranked from the most to the least frequently reported.

#### Disease modifiers

To identify demographic characteristics, comorbidities and pre-conditions associated with COVID-19 symptom severity in pregnancy, a multivariate unconditional regression was applied to each dataset, with the severity index as a response variable and age, diabetes, heart, lung (and asthma) and kidney diseases as factors. As the regression investigated within-group factors, age-weighting was not applied. Bonferroni correction for multiple tests was applied.

Statistical analyses were performed using STATA version 16 (discovery cohort) and R 3.6.3 (replication cohort).

## 3. Results

### Cohort Characteristics and COVID-19 Outcomes

The discovery cohort (N=400,750 participants) was obtained from women (aged 18-44) in the test subset only. It includes longitudinal records from 14,049 pregnant and 386,701 non-pregnant women who had a median duration of follow-up of 18 days (IQR [6-34]) and contributed to an average of 6.6 reports per woman. Among the 45% of pregnant women who self-reported their gestation week at baseline, 14% were in the first trimester, 43% were in the second trimester, and 43% were in the third trimester. The replication cohort consisted of N= 1,344,966 cross-sectional surveys from women aged 18-44. One-time surveys were administered over the 9 week period, at average rate of about 149 thousand surveys per week, using survey methodology. There were 41,796 surveys from women who indicated they were pregnant (3.1% of the source population). Demography was consistent with US age-specific pregnancy rates and stable over the survey period ^23^.

Demographic details are shown in Table 1, together with testing rates. In the discovery cohort, we identified 629 and 25,061 pregnant and non-pregnant women, respectively, who were suspected positive for SARS-CoV-2 infection based on the symptom-score-based imputation method. Of these suspected positive, 21 (3.3%) pregnant and 591 (2.4%) non-pregnant were hospitalized, respectively. In the replication cohort, the proportion of 1,076 (2.9%) suspected positive pregnant was slightly lower compared to 44,772 (4.0%) suspected positive non-pregnant.

**Table 1.** Characteristics of the two cohorts, presented as percentages and means (standard deviations) in the cohorts. Except for group age, percentages and means are age standardized to the pregnant population age distribution in each cohort. Adjustment for survey weights was applied to the replication cohort. Self-report of being seen at a hospital was used as a proxy for hospitalization in the replication cohort.

|  | Discovery Cohort |  |  | Replication Cohort |  |  |
| --- | --- | --- | --- | --- | --- | --- |
|  | All<br>(N=400,<br>750) | Non-pregnant<br>(N=386,70<br>1) | Pregnant<br>(N=14,04<br>9) | All<br>(N=1,34<br>4,966) | Non-pregnant<br>(N=1,303,1<br>70) | Pregnant<br>(N=41,796) |
| Age (years)<br>(not age-standardized) | 32.1<br>(7.2) | 32.1 (7.3) | 32.4 (4.9) | 29.0<br>(0.02) | 29.0 (0.01) | 29.0 (0.05) |
| Tested | 7.0% | 6.1% | 8.0% | 2.5% | 2.4% | 2.7% |
| Positive | 0.6% | 0.7% | 0.6% | 0.4% | 0.4% | 0.4% |
| Negative | 5.5% | 4.9% | 6.2% | 2.2% | 2.1% | 2.2% |
| Suspected | 5.6% | 6.7% | 4.5% | 3.5% | 4.0% | 3.0% |
| Comorbidities |  |  |  |  |  |  |
| Diabetes | 1.8% | 1.2% | 2.3% | 3.9% | 3.5% | 4.3% |
| Lung | 12.9% | 12.8% | 11.3% | 19.3% | 19.8% | 18.8% |
| Heart | 0.6% | 0.5% | 0.6% | 0.8% | 0.9% | 0.7% |
| Kidney | 0.3% | 0.4% | 0.3% | 0.6% | 0.7% | 0.5% |
| Cancer | 0.1% | 0.2% | 0.1% | 0.9% | 1.1% | 0.8% |
| Symptom<br>Severity | 0.07<br>(0.11) | 0.07 (0.11) | 0.04<br>(0.09) | 0.08<br>(0.0005) | 0.08<br>(0.0003) | 0.07 (0.001) |
| Test positive and<br>hospitalized* | 0.09% | 0.07% | 0.1% | 0.06 % | 0.03% | 0.09% |
| Suspected<br>positive and<br>Hospitalized* | 0.16% | 0.16% | 0.15% | 0.17 % | 0.12% | 0.23% |

\* Hospitalization not queried in replication cohort. Proportion of who tested positive or were suspected positive and who reported seeking care at a hospital for symptoms in the prior 24 hours provided as a proxy.

Validation of the imputation method in a subset of the discovery cohort, and in the replication cohort is depicted in Figure 1, with additional sensitivity analyses in Supplementary Material 2.

**Figure 1.**
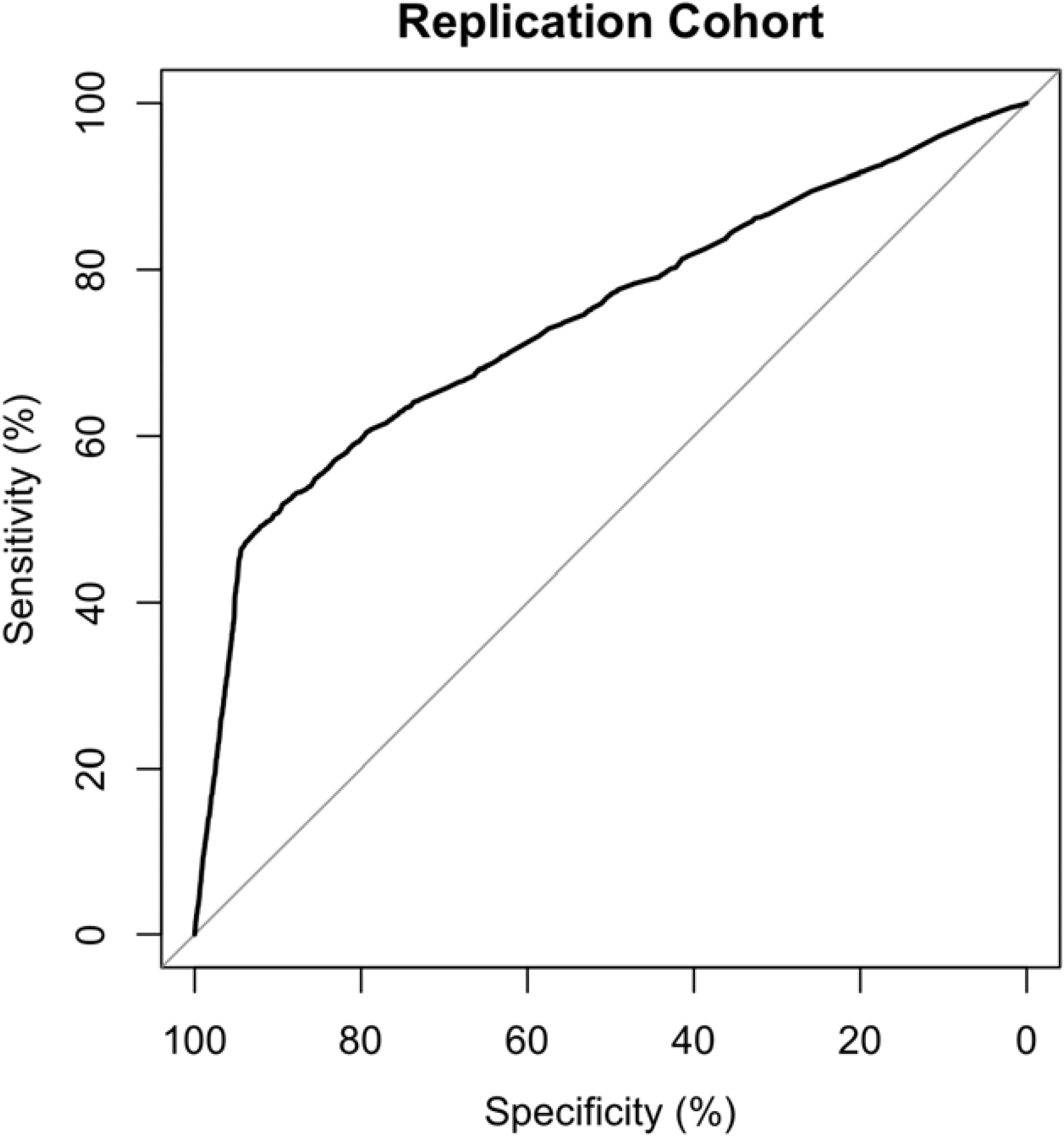
Receiver Operating Characteristics curve showing validation of the imputation of SARS-CoV-2 test status using the mapped symptom score probability in the replication cohort. Area under the curve is 74%.

### Symptomatic, Syndromic and Severity Predictors

Frequency of symptoms and body system clusters is reported in Table 2, and graphically in Figure 2. In the discovery cohort, the most frequent symptoms in the hospitalized pregnant women positive for SARS-CoV-2 were persistent cough, headache and anosmia (all 80.0%), chest pain (73.3%), sore throat and fatigue (66.7%). In the replication cohort, among pregnant test positive women who were seen at the hospital for their illness, the most frequent symptoms were fatigue (87.5%), cough (84.6%), nausea or vomiting (78.2%), muscle pain (76.2) and anosmia (75.2%).

**Table 2.** Frequencies and percentage values of presentation of each symptom among hospitalized in the discovery cohort, and among all women who self-reported being seen at a hospital for their illness in the replication cohort (as hospitalization data were not available). Data are reported by pregnancy status and further subdivided by SARS-CoV-2 test positive or suspected COVID-19 status. Data are reported as N (%) in the discovery cohort, and N surveys (survey-weight adjusted %) in the replication cohort. *Fatigue* was mapped to *tiredness/exhaustion* and *unusual muscle pain* to *pain in muscle and joints* in the replication cohort. Symptoms not ascertained or mapped in either cohort are marked as not available (NA).

| Cluster (body system) | Symptom | Discovery Cohort |  |  |  | Replication Cohort |  |  |  |
| --- | --- | --- | --- | --- | --- | --- | --- | --- | --- |
|  |  | Hospitalised non pregnant positive (N=229) | Hospitalised non pregnant suspected positive (N=591) | Hospitalised pregnant positive (N=15) | Hospitalised pregnant suspected positive (N=21) | Seen at hospital, non-pregnant positive (N=300) | Seen at hospital, non-pregnant suspected positive (N=1395) | Seen at hospital, pregnant positive (N=29) | Seen at hospital, pregnant suspected positive (N= 75) |
| Inflammation | Fever | 151 (65.9) | 359 (60.7) | 8 (53.3) | 12 (57.1) | 135 (48.1) | 514 (39.0) | 12 (50.6) | 19 (29.9) |
|  | Unusual muscle pain | 121 (52.8) | 338 (57.2) | 9 (60.0) | 9 (42.9) | 199 (69.0) | 1,048 (77.0) | 19 (76.2) | 52 (71.8) |
|  | Fatigue | 125 (54.6) | 345 (58.4) | 10 (66.7) | 8 (38.1) | 207 (65.9) | 1,142 (79.8) | 24 (87.5) | 61 (84.0) |
| Neurologic | Headache | 185 (80.8) | 516 (87.3) | 12 (80.0) | 17 (81.0) | NA | NA | NA | NA |
|  | Delirium | 88 (38.4) | 253 (42.8) | 4 (26.7) | 1 (4.8) | NA | NA | NA | NA |
| Cardiopulmonary | Dyspnea | 113 (49.3) | 316 (53.5) | 9 (60.0) | 11 (52.4) | 166 (54.8) | 913(65.1) | 20 (73.6) | 47 (66.9) |
|  | Persistent cough | 178 (77.7) | 438 (74.1) | 12 (80.0) | 19 (90.5) | 202 (68.2) | 1,161 (82.3) | 24 (84.6) | 61 (81.0) |
|  | Chest pain | 170 (74.2) | 463 (78.3) | 11 (73.3) | 14 (66.7) | 156 (53.2) | 787 (56.8) | 17 (62.3) | 34 (51.9) |
|  | Difficulty breathing | NA | NA | NA | NA | 144 (47.7) | 710 (51.6) | 16 (56.0) | 36 (55.1) |
| Oropharyngeal | Hoarse voice | 117 (51.1) | 309 (52.3) | 6 (40.0) | 11 (52.4) | NA | NA | NA | NA |
|  | Sore throat | 148 (64.6) | 371 (62.8) | 10 (66.7) | 14 (66.7) | 118 (38.3) | 552(39.1) | 15 (59.0) | 29 (46.7) |
|  | Nasal congestion | NA | NA | NA | NA | 146 (48.4) | 719 (51.5) | 19 (61.5) | 45 (56.2) |
|  | Runny nose | NA | NA | NA | NA | 116 (35.9) | 636 (48.5) | 14 (57.0) | 33 (51.4) |
| Anosmia/ageusia | Anosmia | 177 (77.3) | 481 (81.4) | 12 (80.0) | 19 (90.5) | 182 (63.1) | 786 (56.7) | 20 (75.2) | 47 (70.4) |
| Gastrointestinal | Skipped meals | 153 (66.8) | 400 (67.7) | 7 (46.7) | 11 (52.4) | NA | NA | NA | NA |
|  | Abdominal pain | 115 (50.2) | 274 (46.4) | 9 (60.0) | 10 (47.6) | NA | NA | NA | NA |
|  | Diarrhoea | 126 (55.0) | 275 (46.5) | 7 (46.7) | 11 (52.4) | 137 (49.2) | 611 (44.4) | 17 (59.8) | 39 (56.1) |
|  | Nausea or vomiting | NA | NA | NA | NA | 138 (49.4) | 633 (49.8) | 21 (78.2) | 51 (79.4) |

**Figure 2.**
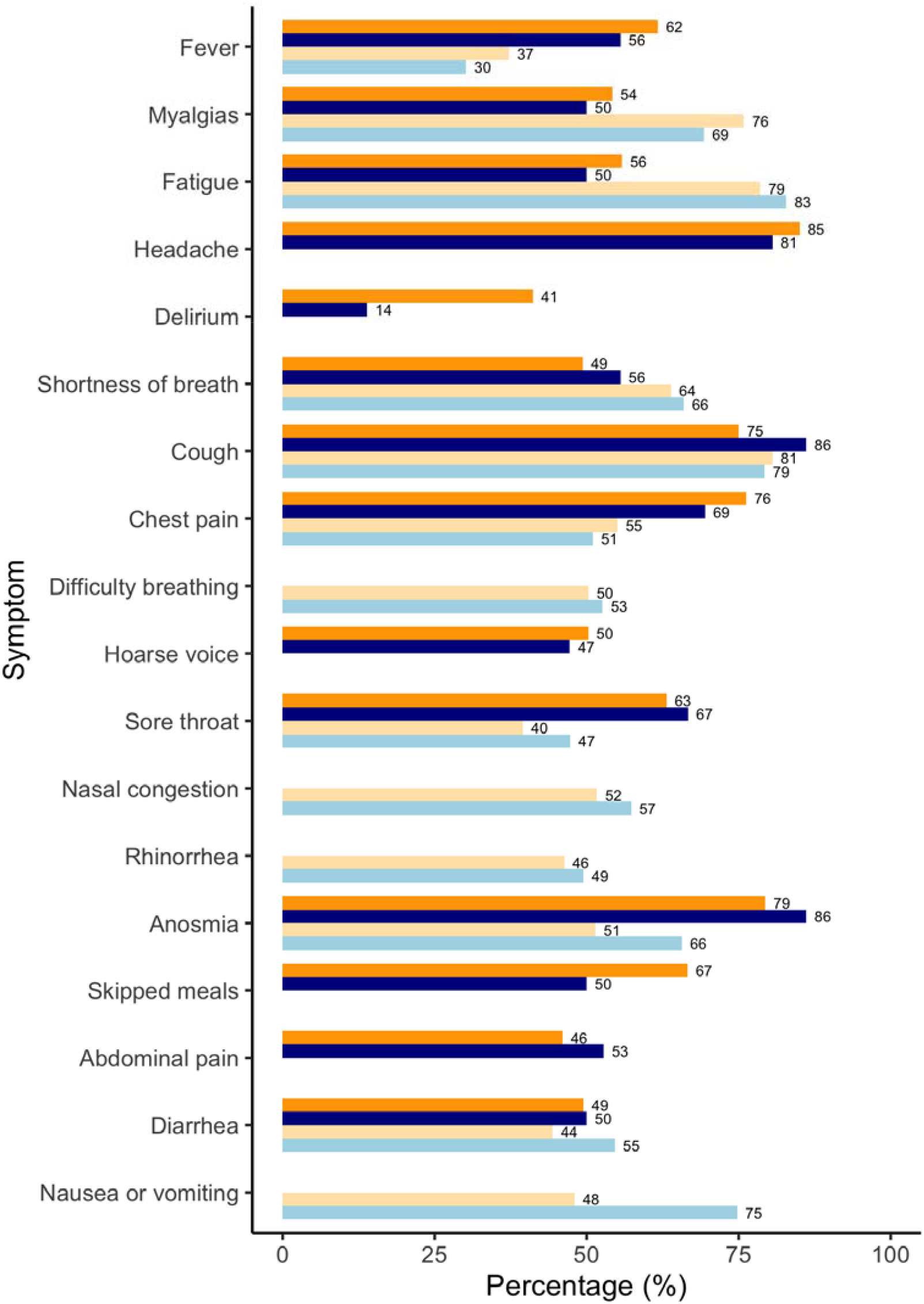
Comparison of symptoms presentation in the discovery (DC) and replication (RC) cohorts. Results refer to non-pregnant (orange) and pregnant (blue) women tested positive and suspected positive for SARS-CoV-2 and who required hospitalization (in DC, darker shade) or were seen at the hospital (RC, lighter shade). Results are reported as age-standardized percentage of women reporting each symptom in each sub-cohort.

In the discovery cohort, univariate analysis on each symptom found significant effect of pregnancy for decreased odds of *skipped meals* (OR 0.5, 95% CI 0.2 to 0.9) and of *delirium* (OR 0.2, 95% CI 0.1 to 0.6) but not for the other symptoms. Multivariate logistic regression found lower frequency of neurologic symptoms (OR 0.3, 95% CI 0.2 to 0.6) for the positive hospitalized pregnant vs. non pregnant women. Among test positives in the replication cohort, pregnancy status was most strongly associated with increased odds of *nausea or vomiting* (OR 2.3, 95% confidence interval 1.5 to 3.5) and the *oropharyngeal* cluster (OR 1.6, 95% CI 1.2 to 2.2), even among test positives reporting being seen at a hospital for their illness (OR 3.4, 95% CI 1.3 to 8.8 and OR 2.1, 95% CI 1.1 to 4.1, respectively), indicating how questions are asked can impact symptom profiles in this population (all age-standardized and p<5e-05 Bonferroni corrected).

Univariate weighted regression also showed that pregnancy had no statistically significant effect on the severity of manifestation of SARS-CoV-2 infection, when expressed as ‘severity index’ in both cohorts (p>0.001, uncorrected to test the null hypothesis). In the discovery cohort, overall duration of disease was similar for pregnant and non-pregnant women. However, time to peak of symptom manifestation was statistically longer in pregnant women (mean time = 2.8 days) than in non-pregnant (2.2 days, p=5.5e-7), though clinically the difference may not be significant. In the replication cohort, pregnant women who tested positive and reported being seen at the hospital similarly reported a longer duration of illness.

As mentioned above, in the discovery cohort hospitalized positive pregnant women manifested persistent cough, headache and anosmia (all 80%), chest pain (73.3%), sore throat and fatigue (66.7%) as the most frequent symptoms. Non-hospitalised pregnant women positive for SARS-CoV-2 reported headache (71.9%), anosmia (62.5%), persistent cough (57.8%) and skipped meals (48.4%) most commonly (Figure 3). See Supplementary Material 6 for full list of symptoms and their associated prevalence.

**Figure 3.**
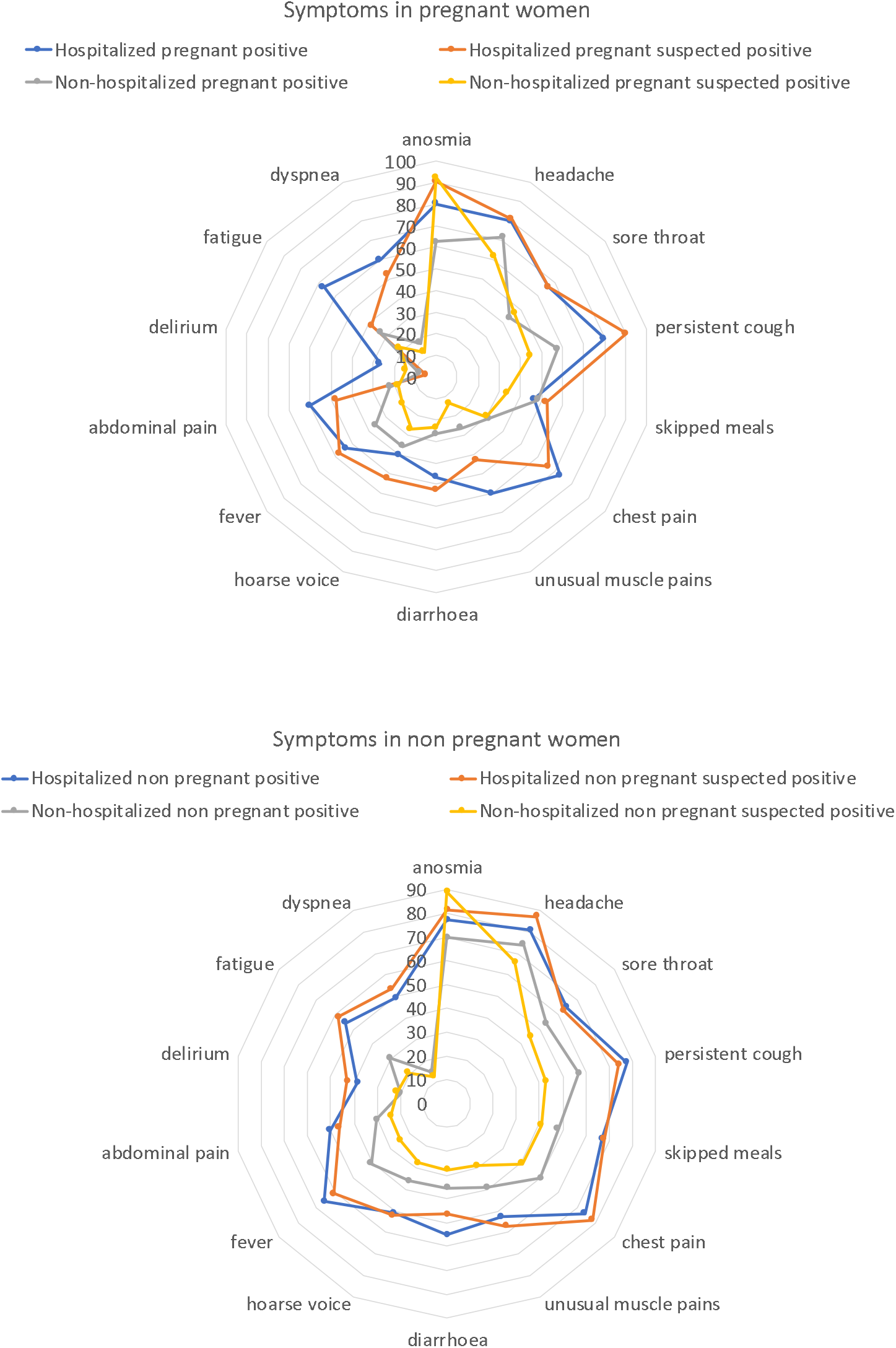
Symptom profile of hospitalized and non-hospitalized pregnant and non-pregnant women positive and suspected positive to SARS-CoV-2 in the discovery cohort. Results are reported in percentage of women reporting each symptom in each group.

### Comorbidities

Lung disease was the comorbidity that most impacted on the severity of symptoms in pregnant positive women (t=4.1 for discovery cohort; t=14.1 for replication cohort, all p-val<0.0001 Bonferroni corrected).

**Table 3.** Frequencies and percentages of comorbidities and pre-existing conditions in the discovery and replication cohorts. Columns refer to pregnant women tested and suspected positive for SARS-CoV-2 infection. Data are reported as N (%). Data from the replication cohort are reported as N surveys (survey-weight adjusted %). Conditions not ascertained or mapped in either cohort are marked as not available (NA).

| Comorbidity or pre-existing condition | Discovery Cohort |  | Replication Cohort |  |
| --- | --- | --- | --- | --- |
|  | Pregnant test positive (N=79) | Pregnant suspected positive (N=629) | Pregnant test positive (N=134) | Pregnant suspected positive (N=1076) |
| Diabetes | 3 (3.8) | 15 (2.4) | 11 (8.9) | 76 (7.4) |
| Lung disease | 8 (10.1) | 80 (12.7) | 37 (31) | 376 (34.2) |
| Heart disease | 1 (1.3) | 5 (0.8) | 5 (6.3) | 41 (4.8) |
| Kidney disease | 0 (0.0) | 2 (0.3) | 8 (7.8) | 30 (43.3) |
| Hypertension | NA | NA | 17 (13.9) | 170 (15.4) |
| Autoimmune | 0 (0.0) | 8 (1.3) | 14 (11.5) | 106 (9.3) |
| Cancer | 0 (0.0) | 1 (0.2) | 5 (4.7) | 29 (3.2) |
| Smoking /<br>Past smoker | 6 (7.6)<br>13 (16.5) | 36 (5.7)<br>121 (19.2) | NA | NA |

In the replication cohort heart disease (t=7.1) also impacted on the severity of symptoms, followed by kidney disease (t=4.6) and diabetes (t=3.6, all significant after Bonferroni correction at p-val<0.0001).

## 4. Discussion

### Main Findings

We studied two large cohorts of women, tested and suspected SARS-CoV-2 positive, with self-reported pregnancy status, symptoms and outcomes through participative surveillance. Pregnant women reported more frequent testing for SARS-CoV-2 than non-pregnant women, but generally did not experience more severe disease. Disease trajectories were similar, and the time from onset to peak of symptoms was only slightly longer in pregnant than non-pregnant women (2.8 vs. 2.2 days).

Gastrointestinal symptoms were different in pregnant and non-pregnant women with poor outcomes, with decreased *skipped meals* in the discovery cohort and increased *nausea or vomiting* in the replication cohort. Neurologic symptoms (only surveyed in the discovery cohort) were decreased in pregnant women.

The current epidemiologic literature is largely based on pregnant women admitted to the hospital, which provides a narrow view of the spectrum of SARS-CoV-2 infection in all pregnant women. Our data show the preponderance of tested positive and even suspected positive pregnant women were not seen at or admitted to the hospital for their illness; most pregnant women reported they recover in the community, as was observed by Lokken et al. ^24^. Cardiopulmonary symptoms were more frequently reported by pregnant women who were hospitalised. Notably, pre-existing lung disease was confirmed as the largest risk factor to develop more severe COVID-19 symptoms in pregnancy, as it is outside of pregnancy. Heart disease, kidney disease and diabetes were also risk factors.

### Interpretation

Pregnant women are considered a high-risk group in UK and were considered high risk in the USA early in the pandemic. This likely contributed to the higher testing proportion but lower positives results among pregnant women vs. non-pregnant. Hospitalized pregnant women presented lower frequency of neurologic symptoms, especially *delirium*, which were only measured in the discovery cohort. The replication cohort showed higher frequency of *gastrointestinal* symptoms among pregnant women with more severe outcomes, especially *nausea or vomiting* in pregnancy, which may be a feature of pregnancy itself (e.g. hyperemesis gravidum). Diarrhoea in positive pregnant women has been previously reported (rates between 8.8% and 14%) ^25,26^.

Syndrome severity did not differ between pregnant and non-pregnant women in both datasets. This posits an equivalent manifestation of SARS-CoV-2 infection in pregnant and non-pregnant, as already reported by Chen and others ^9,12^.

Pre-existing lung disease is the comorbidity with strongest impact on the SARS-CoV-2 infection severity in pregnant women in both cohorts. Lokken et al. ^24^ similarly reported asthma as a primary risk factor for severe COVID-19 in pregnancy. Heart disease, kidney diseases and diabetes were also associated with severity in the replication cohort which had high enough prevalence of these conditions (related to survey-sampling to the general population) to detect an effect. These comorbidities are consistent with risk factors in the general, non-pregnant population; Li et al. observed chronic obstructive pulmonary disease, diabetes, hypertension, coronary heart disease and cerebrovascular diseases had the highest odd ratio for SARS-CoV-2 and admission to the intensive care unit (ICU) ^27^, while Kumar et al. found diabetes increased SARS-CoV-2 severity and mortality two-fold ^28^.

Cough, chest pain and dyspnea showed much higher incidence in the hospitalized pregnant women, indicating that cardiopulmonary symptoms are the major discriminant for hospitalization. Similarly, Ellington et al ^29^, found increased ICU admissions and need of mechanical ventilation in pregnant women, although the cohort studied had higher frequency of underlying medical conditions, and might be less representative of the general pregnant population.

Pregnant women with pre-existing lung disease or prominent cardiopulmonary symptoms may need special attention during the COVID-19 pandemic; lung disease had strongest impact on syndrome severity while cardiopulmonary symptoms were the main factor predicting hospitalization in pregnancy. Indeed, in pregnancy, cardiopulmonary reserve is limited which increases morbidity and complicates management when there are added physiologic stressors (e.g. asthma exacerbation) ^30–32 33^. Diabetes was more common in the pregnant women in our cohorts, likely related to gestational diabetes. We confirmed diabetes is associated to increased severity of SARS-CoV-2 symptoms ^34^.

This study leveraged two cohorts followed through remote, participatory epidemiology, enabling rapid assessment of COVID-19 in pregnancy. The longitudinal nature of the discovery dataset enabled the comparison of disease duration, time from onset to peak of symptoms, and hospitalization between pregnant and non-pregnant women, prospectively. Broadly, pregnancy does not substantially contribute to morbidity in our community-based cohorts. Clinicians should be more vigilant with pregnant who have pre-existing health conditions, prominent respiratory symptoms or a higher severity index -- as is the case in the general population. Further studies specifically targeting high-risk pregnancies and outcomes across the three trimesters may be warranted, to better define outcomes in this population. Also, we point out the need to interpret hospitalization rates and severity results in light of the policy changes, which can be dependent on the context or country.

### Strengths and limitations

Participatory surveillance tools are crucial to epidemiological research and citizen science, as they increase population’s awareness of urgent public health risks, promote public participation into science and enable inclusion in studies of large samples from the community within short time periods. Real-time public health data has been crucial in decision-making during the COVID-19 pandemic. However, user of smartphone applications and web-based surveys may not be representative of the general population, potentially limiting generalizability. Self-reported events may suffer from misclassification bias, which may be differential (e.g. ability to log hospitalization may be higher in less severely affected participants, test results known at the time of cross-sectional symptom reporting may differ). Median app usage was 18 days, which may be insufficient follow-up to ascertain all outcomes. In the discovery cohort, pregnancy status was only queried at the time of registration; women who became pregnant after registration may be misclassified. In addition, gestational age during the infection could not be assessed, as well as whether women were symptomatic at the time of delivery. The replication cohort was designed to be representative of USA population through survey sampling for the active user base and weights with raking to the USA census. Despite the different platforms and country of origin of users, the cross-sectional surveys showed similar results to the detailed longitudinal discovery cohort of technology-aware smartphone users. However, it was not possible to distinguish difference in methodology from country-specific effects. Additionally, we applied age-standardization to account for demographic structure inherent to pregnancy. Despite the differences in the UK, USA and Sweden testing guidelines and healthcare systems, morbidity with COVID-19 in pregnancy were comparable. We were able to develop and validate a prediction score for suspected positive, as well as a severity score for use in women of childbearing age, and these performed similarly in the cross-sectional survey data despite development using longitudinal symptom reports. This may be useful for obstetricians in the context of limited access to SARS-CoV-2 testing during this pandemic.

### Conclusions

Our findings from two large real-time syndromic surveillance technologies provide evidence that most pregnant women in the community who are positive for SARS-CoV-2 are at similar risk of developing either increased morbidity or complex symptomatology compared with non-pregnant women. However, pre-existing lung or cardiac disease may exacerbate cardiopulmonary stress of pregnancy. Pregnant women with comorbidities appear to be at increased risk for severe disease, consistent with evidence from COVID-19 infection in the general population. Pregnant women with pre-existing conditions, similar to the general adult population, require careful monitoring for the evolution of their symptoms during SARS-CoV-2 infection.

## Data Availability

Data are available.

https://covid.joinzoe.com/data

## Acknowledgements

Authors express gratitude to all the participants who entered data into the smartphone app and website, including study volunteers enrolled in the Coronavirus Pandemic Epidemiology (COPE) consortium and Carnegie Mellon Delphi Research Center. We thank the staff of Zoe Global, the Department of Twin Research at King’s College London, the Clinical and Translational Epidemiology Unit at Massachusetts General Hospital, the Department of Clinical Sciences in Malmö at Lund University and the Department of Medical Sciences at Uppsala University for tireless work in contributing to the running of the study and data collection.

## References

1. Gorbalenya AE, Baker SC, Baric RS, de Groot RJ, Drosten C, Gulyaeva AA, et al. The species Severe acute respiratory syndrome-related coronavirus: classifying 2019-nCoV and naming it SARS-CoV-2. Nature Microbiology. 2020.

2. Sironi M, Hasnain SE, Rosenthal B, Phan T, Luciani F, Shaw MA, et al. SARS-CoV-2 and COVID-19: A genetic, epidemiological, and evolutionary perspective. Infection, Genetics and Evolution. 2020.

3. Wong SF, Chow KM, Leung TN, Ng WF, Ng TK, Shek CC, et al. Pregnancy and perinatal outcomes of women with severe acute respiratory syndrome. Am J Obstet Gynecol. 2004;

4. Park MH, Kim HR, Choi DH, Sung JH, Kim JH. Emergency cesarean section in an epidemic of the middle east respiratory syndrome: A case report. Korean J Anesthesiol. 2016;

5. Chui ML, Shell FW, Tse NL, Kam MC, Wai CY, Tin YW, et al. A case-controlled study comparing clinical course and outcomes of pregnant and non-pregnant women with severe acute respiratory syndrome. BJOG An Int J Obstet Gynaecol. 2004;

6. Panahi L, Amiri M, Pouy S. Risks of Novel Coronavirus Disease (COVID-19) in Pregnancy; a Narrative Review. Arch Acad Emerg Med. 2020;

7. Khalil A, Kalafata E, Benlioglua C, O’Brien P, Morris E, Draycott T, et al. SARS-CoV-2 infection in pregnancy: A systematic review and meta-analysis of clinical features and pregnancy outcomes. EClinicalMedicine. 2020;100446.

8. Khalil A, Von Dadelszen P, Draycott T, Ugwumadu A, Magee LA. Increase in the incidence of stillbirth during the COVID-19 pandemic. JAMA - J Am Med Assoc. 2020;July 10.

9. Chen Y, Li Z, Zhang YY, Zhao WH, Yu ZY. Maternal health care management during the outbreak of coronavirus disease 2019. Journal of Medical Virology. 2020.

10. CDC Coronavirus Disease 2019 (COVID-19) People Who Need Extra Precautions Others At Risk. If You Are Pregnant, Breastfeeding, or Caring for Young Children. 2020.

11. Hu Y, Sun J, Dai Z, Deng H, Li X, Huang Q, et al. Prevalence and severity of corona virus disease 2019 (COVID-19): A systematic review and meta-analysis. Journal of Clinical Virology. 2020.

12. Knight M, Bunch K, Vousden N, Morris E, Simpson N, Gale C, et al. Characteristics and outcomes of pregnant women admitted to hospital with confirmed SARS-CoV-2 infection in UK: national population based cohort study. BMJ. 2020;

13. Chen H, Guo J, Wang C, Luo F, Yu X, Zhang W, et al. Clinical characteristics and intrauterine vertical transmission potential of COVID-19 infection in nine pregnant women: a retrospective review of medical records. Lancet. 2020;

14. Ceulemans M, Verbakel JY, Van Calsteren K, Eerdekens A, Allegaert K, Foulon V. SARS-CoV-2 Infections and Impact of the COVID-19 Pandemic in Pregnancy and Breastfeeding: Results from an Observational Study in Primary Care in Belgium. Int J Env Res Public Heal. 2020;17(18):E6766.

15. McCullough Pa, Eidt J, Rangaswami J, Lerma E, Tumlin J, Wheelan K, et al. Urgent need for individual mobile phone and institutional reporting of at home, hospitalized, and intensive care unit cases of SARS-CoV-2 (COVID-19) infection. Reviews in cardiovascular medicine. 2020.

16. Brownstein JS, Freifeld CC, Madoff LC. Digital disease detection - Harnessing the web for public health surveillance. New England Journal of Medicine. 2009.

17. Hussain T, Smith P, Yee LM. Mobile Phone-Based Behavioral Interventions in Pregnancy to Promote Maternal and Fetal Health in High-Income Countries: Systematic Review. JMIR mHealth and uHealth. 2020.

18. mHealth solutions list [Internet]. p. http://mhealth-hub.org/mhealth-solutions-against-c. Available from: http://mhealth-hub.org/mhealth-solutions-against-covid-19

19. Drew DA, Nguyen LH, Steves CJ, Menni C, Freydin M, Varsavsky T, et al. Rapid implementation of mobile technology for real-time epidemiology of COVID-19. Science (80-). 2020;

20. Menni C, Valdes AM, Freidin MB, Sudre CH, Nguyen LH, Drew DA, et al. Real-time tracking of self-reported symptoms to predict potential COVID-19. Nat Med. 2020;

21. Facebook Questionnaire [Internet]. p. https://cmu.ca1.qualtrics.com/jfe/preview/SV_cT2ri. Available from: https://cmu.ca1.qualtrics.com/jfe/preview/SV_cT2ri3tFp2dhJGZ?Q_SurveyVersionID=curre nt&Q_CHL=preview

22. Kreuter F, Barkay N, Bilinski A, Bradford A, Chiu S, Eliat R, et al. Partnering with Facebook on a university-based rapid turn-around global survey. Surv Res Methods. 2020;14(2).

23. ZCDC D. https://www.cdc.gov/nchs/data/databriefs/db136.pdf [Internet]. CDC data. 2020. Available from: https://www.cdc.gov/nchs/data/databriefs/db136.pdf

24. Lokken EM, Walker CL, Delaney S, Kachikis A, Kretzer NM, Erickson A, et al. Clinical Characteristics of 46 Pregnant Women with a SARS-CoV-2 Infection in Washington State. Am J Obstet Gynecol. 2020;

25. Yu N, Li W, Kang Q, Xiong Z, Wang S, Lin X, et al. Clinical features and obstetric and neonatal outcomes of pregnant patients with COVID-19 in Wuhan, China: a retrospective, single-centre, descriptive study. Lancet Infect Dis. 2020;

26. Yang Z, Wang M, Zhu Z, Liu Y. Coronavirus Disease 2019 (COVID-19) and Pregnancy: A Systematic Review. J Matern Fetal Neonatal Med. 2020;Apr 30:1–4.

27. Li J, Xue H, Yuan YUAN, Wei Z, Li X, Zhang Y, et al. Meta-analysis Investigating the Relationship Between Clinical Features, Outcomes, and Severity of Severe Acute Respiratory Syndrome Coronavirus 2 (SARS-CoV-2) Pneumonia. Am J Infect Control. 2020;Jun 12(S0196-6553(20)30369-2).

28. Kumar A, Arora A, Sharma P, Anikhindi SA, Bansal N, Singla V, et al. Is diabetes mellitus associated with mortality and severity of COVID-19? A meta-analysis. Diabetes Metab Syndr Clin Res Rev. 2020;

29. Ellington S, Strid P, Tong VT, Woodworth K, Galang RR, Zambrano LD, et al.Characteristics of Women of Reproductive Age with Laboratory-Confirmed SARS-CoV-2 Infection by Pregnancy Status — United States, January 22–June 7, 2020. MMWR Morb Mortal Wkly Rep. 2020;

30. Li N, Han L, Peng M, Lv Y, Ouyang Y, Liu K, et al. Maternal and neonatal outcomes of pregnant women with COVID-19 pneumonia: a case-control study. Clin Infect Dis. 2020;

31. Qiao J. What are the risks of COVID-19 infection in pregnant women? The Lancet. 2020.

32. Gardner MO, Doyle NM. Asthma in pregnancy. Obstetrics and Gynecology Clinics of North America. 2004.

33. Angeli F, Spanevello A, De Ponti R, Visca D, Marazzato J, Palmiotto G, et al.Electrocardiographic features of patients with COVID-19 pneumonia. Eur J Intern Med. 2020;

34. Kayem G, Lecarpentier E, Deruelle P, Bretelle F, Schmitz T, Alessandrini V, et al. A snapshot of the Covid-19 pandemic among pregnant women in France. J Gynecol Obstet Hum Reprod. 2020;

